# Increased Detection coupled with Social Distancing and Health Capacity Planning Reduce the Burden of COVID-19 Cases and Fatalities: A Proof of Concept Study using a Stochastic Computational Simulation Model

**DOI:** 10.1101/2020.04.05.20054775

**Authors:** Pramit Ghosh, Salah Basheer, Sandip Paul, Partha Chakrabarti, Jit Sarkar

## Abstract

**Objective:** In absence of any vaccine, the Corona Virus Disease 2019 (COVID-19) pandemic is being contained through a non-pharmaceutical measure termed Social Distancing (SD). However, whether SD alone is enough to flatten the epidemic curve is debatable. Using a Stochastic Computational Simulation Model, we investigated the impact of increasing SD, hospital beds and COVID-19 detection rates in preventing COVID-19 cases and fatalities.

**Research Design and Methods:** The Stochastic Simulation Model was built using the *EpiModel* package in R. As a proof of concept study, we ran the simulation on Kasaragod, the most affected district in Kerala. We added 3 compartments to the SEIR model to obtain a SEIQHRF (Susceptible-Exposed-Infectious-Quarantined-Hospitalised-Recovered-Fatal) model.

**Results:** Implementing SD only delayed the appearance of peak prevalence of COVID-19 cases. Doubling of hospital beds couldn’t reduce the fatal cases probably due to its overwhelming number compared to the hospital beds. Increasing detection rates could significantly flatten the curve and reduce the peak prevalence of cases (increasing detection rate by 5 times could reduce case number to half).

**Conclusions:** An effective strategy to contain the epidemic spread of COVID-19 in India is to increase detection rates in combination with SD measures and increase in hospital beds.

**HIGHLIGHTS:**

- Increased Detection of COVID-19 cases must accompany Social Distancing and Health Capacity Planning to reduce the burden of cases and fatalities.
- Interruptive Social Distancing is an effective alternative to continuous Social Distancing.
- Given the overwhelming burden of COVID-19 fatalities, there is immediate need of co-ordination with the Private Healthcare Sector.
- COVID-19 cases will be peaking after May, 2020 giving us time for Healthcare Capacity Building in the government and private sector both.

## 1. INTRODUCTION

Coronavirus Disease 2019 (COVID-19) caused by the novel corona virus SARS-CoV-2 has been declared a pandemic by World Health Organization [1]. The disease has rapidly spread globally, the first case in India being detected on 30^th^ January, 2020. Since then, several research groups have been working in predicting COVID-19 cases and their consequences in India. Till date, Susceptible-Infectious-Recovered (SIR) models has been widely used in predicting the course of epidemics [2]. SIR models are epidemic simulation models which divide the population into 3 compartments of Susceptible, Infectious and Recovered groups and assume a defined rate of transition between them. Variation of SIR model is the SEIR model which includes another compartment “Exposed” in the model [3].

According to the Report of the WHO-China Joint Mission on Coronavirus Disease 2019 (COVID-19), COVID-19 manifests in several forms with 80% of the cases presenting with mild to moderate disease. 13.8% progress to severe disease and 6.1% become critically ill with respiratory failure, septic shock or multi-organ failure [1]. However, detection and response capacity of COVID-19 cases globally vary across all countries [4]. In a country like India with a varied administrative set-up and state policies, healthcare amenities vary even further [5]. Amidst measures like “*Lock-down*”, Social distancing and Sanitization being used to reduce the peak number of COVID-19 cases [6] in India, we extended the SIR model to a SEIQHRF (Susceptible-Exposed-Infectious-Quarantined-Hospitalised-Recovered-Fatal) model [7] to take in account the healthcare capacity of that state. As a proof of concept study, we ran the SEIQHRF model on Kasargod [8], the most affected district in Kerala, the state with best healthcare capacity in India [9].

## 2. MATERIALS AND METHODS

### 2.1 The Model and its parameters

The Epidemic Model was built using *EpiModel* package [10] in R Language for Statistical Computing. Here we used a stochastic, discrete time, individual contact model so that we could introduce the effects of interventions like “*lock-down*” and social distancing. The model had 7 compartments (Figure1) as follows:

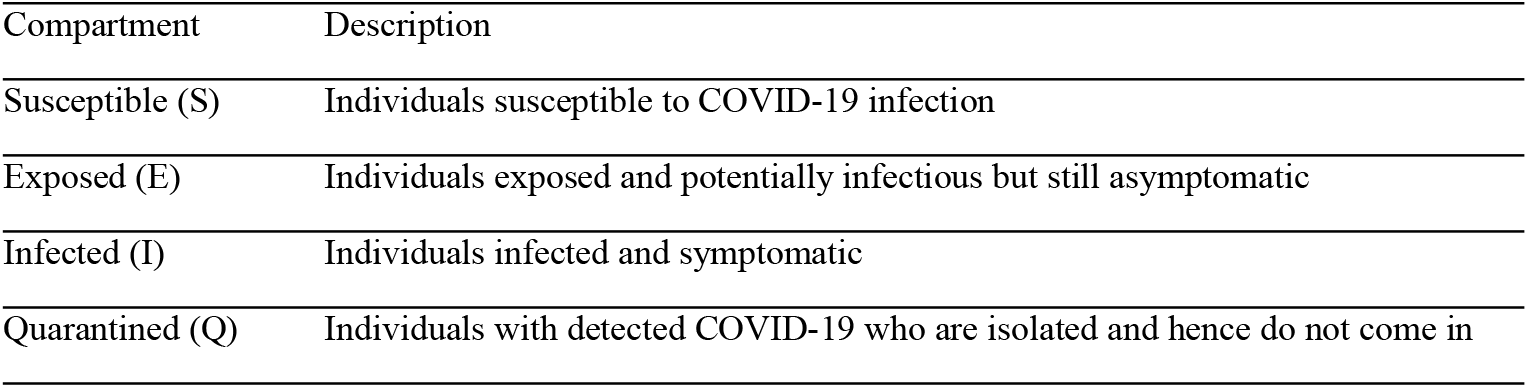
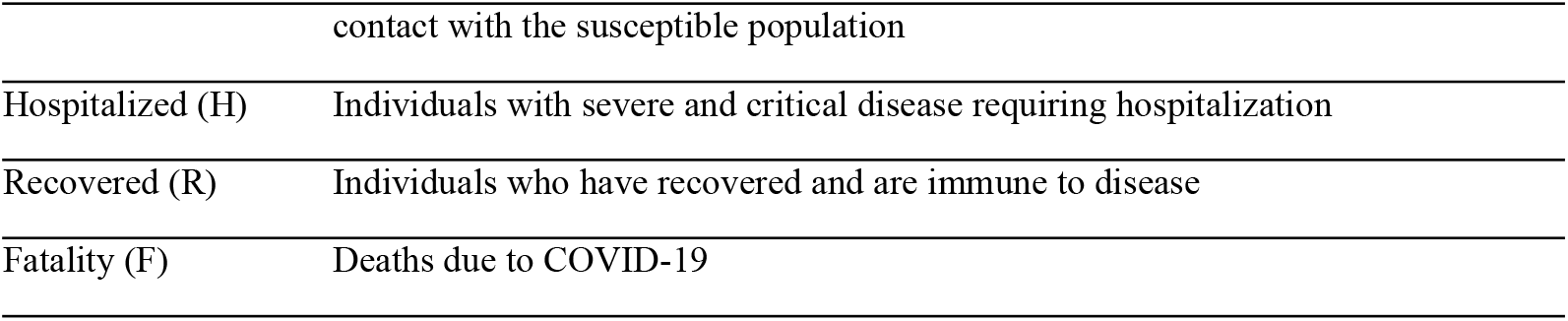

| Compartment | Description |
| --- | --- |
| Susceptible (S) | Individuals susceptible to COVID-19 infection |
| Exposed (E) | Individuals exposed and potentially infectious but still asymptomatic |
| Infected (I) | Individuals infected and symptomatic |
| Quarantined (Q) | Individuals with detected COVID-19 who are isolated and hence do not come in |
|  | contact with the susceptible population |
| Hospitalized (H) | Individuals with severe and critical disease requiring hospitalization |
| Recovered (R) | Individuals who have recovered and are immune to disease |
| Fatality (F) | Deaths due to COVID-19 |

**Figure1:**
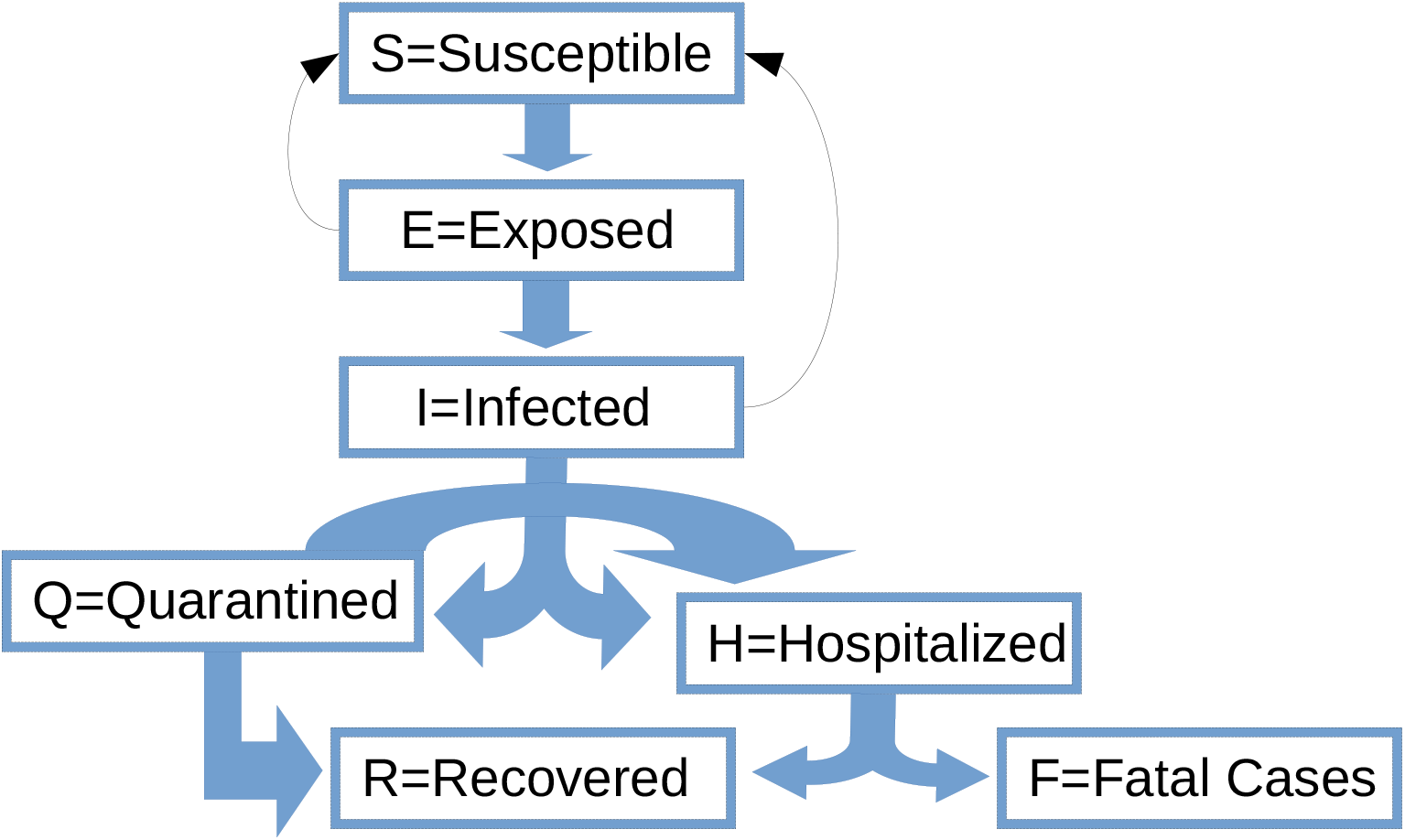
The SEIQHRF Model Compartments in the population. The thick blue arrows show the direction of flow of individuals between compartments. The thin black arrows show the direction of infection through contact from the Infected and Exposed population to the Susceptible population

The following values for the parameters are used in the model-

1. Progression from E to I was determined by random draws from a Weibull distribution with the shape parameter of 1.5 and scale parameter of 5.
2. Transition from I to Q was done with a random sample with a sample fraction of 1/30 (i.e. 1 out of 30 infected people showing symptoms were quarantined daily). Such a low number was considered due to low awareness and readiness to report to the hospital on developing symptoms.
3. Transition from I or Q to H was also done with a random sample with a sample fraction of 1/50 (assuming 20% of the symptomatic cases progress to severe or critical disease with an average illness duration of 10 days).
4. Transition from Q or H to R was also done from a Weibull distribution with the shape parameter of 1.5 and scale parameter of 35.
5. Transition from H to F is determined by a random sample with a sample fraction of 1/50 (considering a baseline mortality of 2% per day for people needing hospitalization). However, depending on the availability of hospital beds, the fatality rate for those who require hospitalization but cannot be hospitalized due to the lack of beds is considered to be 1/25 (double the baseline mortality rate). A fatality time coefficient is considered which linearly increases the fatality rate according to the number of days spent in the H compartment. Use of time coefficient better approximates the trapezoid survival time distribution observed in critical patients [11]. A fatality time coefficient of 0.5 is assumed. As the most optimistic solution, the number of beds available for hospitalization is assumed to be all the beds in the Medicine Department of District Hospitals under the Directorate of Health Services, Government of Kerala [8].
6. Transition from H to R is also a random sample with a sample fraction of 1/15 (considering an average recovery time of 15 days for a patient).
7. The number of exposure events between infectious individuals in I compartment and susceptible individuals in the S compartment was assumed to be 10 (a parameter which can be reduced through Social distancing) and the probability of passing the infection (can be reduced by increasing hygiene measures) was taken as 0.05.
8. Values of exposure events and probability of passing infection from asymptomatic individuals of Exposed (E) compartment to Susceptible population was taken to be 10 and 0.02 (as the duration of infectivity was less in the E compartment compared to the I compartment). The incubation period for COVID-19 is 5.2 days [12] and a recent study has shown the median serial interval to be 4.6 days [13] pointing out to the possibility that presymptomatic transmission is possible for COVID-19.
9. Arrival rate in the model was taken to be (14.61/1000)/365 considering an annual birth rate of 14.61 births/1000 population. Departure rate from all the compartments except the H compartment was taken as (5.44/1000)/365 considering an annual death rate of 5.44 deaths/1000 population [14].
10. The starting day for the simulation was taken as 16^th^ March 2020. The initial susceptible population in Kasargod district was taken as 13,07,375 which was the population in 2011 census [14]. As the incubation period of COVID-19 is 5.2 days (rounding off to 5 days), the number of infected cases (I compartment) on 16^th^ March was assumed to be the number of new cases on 21^st^ March which was 6.
11. Taking all the aforesaid parameters, we ran 8 simulations over 100 days starting from 16^th^ March, 2020 and expressed the results as the mean of all the runs.

### 2.2 Adding Interventions to the model

Like all other epidemics, the COVID-19 epidemic is being intervened across the globe by non-pharmaceutical modes like Social distancing along with increasing the bed capacity in the hospitals. We introduced these 4 measures by modifying the following parameters in the model:

1. Social Distancing starting from 25^th^ March, 2020: The Government of India announced a lock-down from 25^th^ March, 2020 till 14^th^ April, 2020 as an intervention to stop the COVID-19 pandemic [15]. Studies have suggested 25% of the contacts to take place in the workplaces [16]. Hence, as a reflection of Social Distancing, we gradually reduced the number of exposure events from 10 to 5 (being more optimistic) from 25^th^ March to 14^th^ April, 2020 in a time-dependent manner and again increased it back to its initial rate from 15^th^ April, 2020.
2. Increasing Hospital Bed Capacity from 25^th^ March to 14^th^ April, 2020: Having introduced Social Distancing till 14^th^ April, 2020, we also introduced another intervention by increasing the hospital bed capacity to double the initial number considered. As increasing bed capacity is a time-dependent process, we gradually increased the number of beds with time during the same duration when “lock-down” was present i.e. from 25^th^ March, 2020 to 14^th^ April, 2020
3. Our third intervention was to continue the lock-down for another 2 weeks till 28^th^ April, 2020 with a similar increase in bed capacity as mentioned earlier. In this case the number of exposure events at the on 28^th^ April was considered to be 2.5 considering the prolonged duration of awareness and practice on Social distancing.
4. The fourth intervention was to release lock-down for 1 week after 14^th^ April for 7 days and then again continue it for another 21 days till 12^th^ May, 2020 with a similar increase in bed capacity as mentioned earlier. In this case for the second duration of social distancing, it was considered that the number of exposure events will linearly fall from 10 to 2.5 as a consequence of prolonged awareness and readiness of the population.
5. The fifth intervention was to increase the case detection rate by 3 times i.e. transition from I to Q from 1/30 to 1/10 during the first 21 days of Social Distancing and continue the case detection and isolation at that rate.
6. The sixth intervention was to increase the case detection rate by 5 times i.e. transition from I to Q from 1/30 to 1/6 during the first 21 days of Social Distancing and continue the case detection and isolation at that rate.
7. The seventh intervention was to increase the case detection rate by 5 times i.e. transition from I to Q from 1/30 to 1/6 during the first 21 days of Social Distancing and continue Social Distancing for another 14 days.
8. The eight intervention was a combination of increased detection and isolation rate by 5 times i.e. transition from I to Q from 1/30 to 1/6 during the first 21 days of Social Distancing in addition to the fourth intervention.

## 3. RESULTS

### 3.1 Impact of Social Distancing and Increasing Hospital Beds for COVID-19 Infection and Fatality

The baseline simulation shows the prevalence of cases in all the seven compartments over 100 days since 16^th^ March 2020. Expectedly, Susceptible (S) and Recovered (R) cases follow reciprocal distributions and the peak of the Exposed (E) appear before the Infectious (I) compartment followed by Hospitalised (H) and Quarantined (Q) compartments (Figure 2A). Interestingly, the Infectious (I) and Fatal (F) cases reach respective peaks after a lag of 50 days and decline in Hospitalisation (H) with stabilization of Fatal (F) cases appear after 70 days (Figure 2A, B).

**Figure 2:**
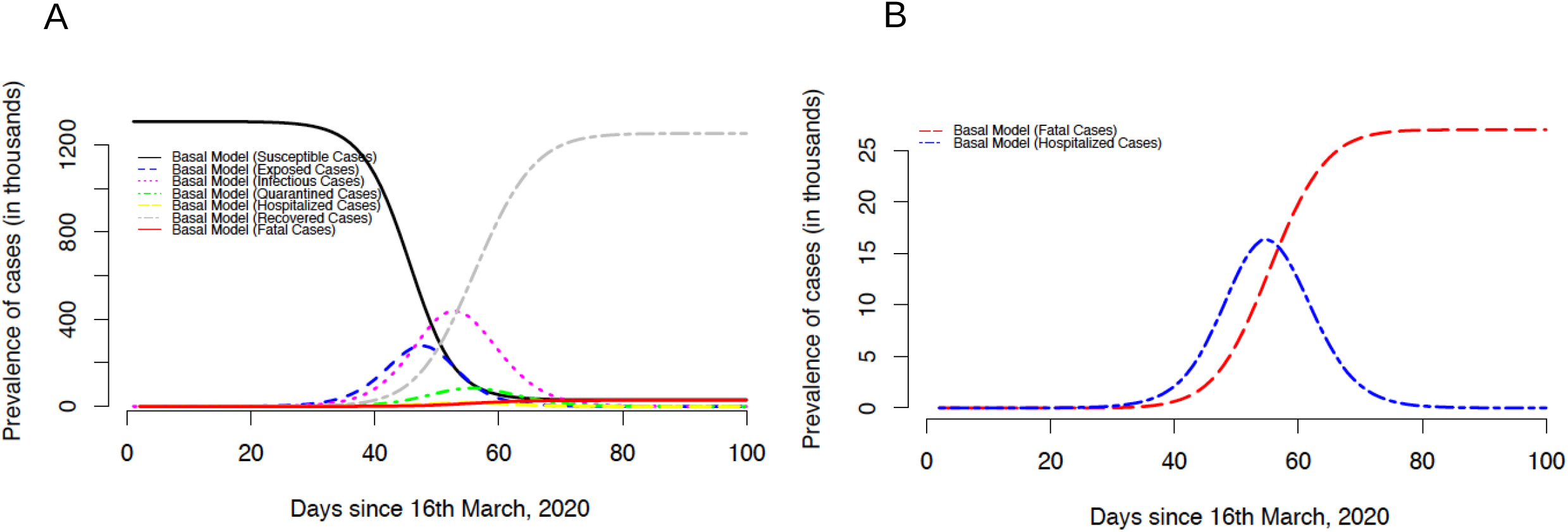
Prevalence of cases in the seven compartments in the baseline SEIQHRF simulation model.

The impact of different Social Distancing (SD) interventions over 3 weeks on the Infectious (I) and Fatal (F) compartments are obtained by running multiple simulations through SD. Interestingly, SD could not decrease the peak prevalence of the I and F compartment but rather delayed them by ∼10 days when the rate of detection and isolation of confirmed cases are kept at the initial value in the baseline model i.e. daily quarantine of 1 in 30 cases (Figure 3A). Extending the duration of SD to a continuous of 5 weeks also delayed the peak prevalence without showing obvious impact on I and F compartments. Surprisingly, doubling the hospital bed capacity had little impact of the peak prevalence of the I and F compartments (Figure 3B), probably due to the overwhelming number of patients compared to the bed capacity.

**Figure 3:**
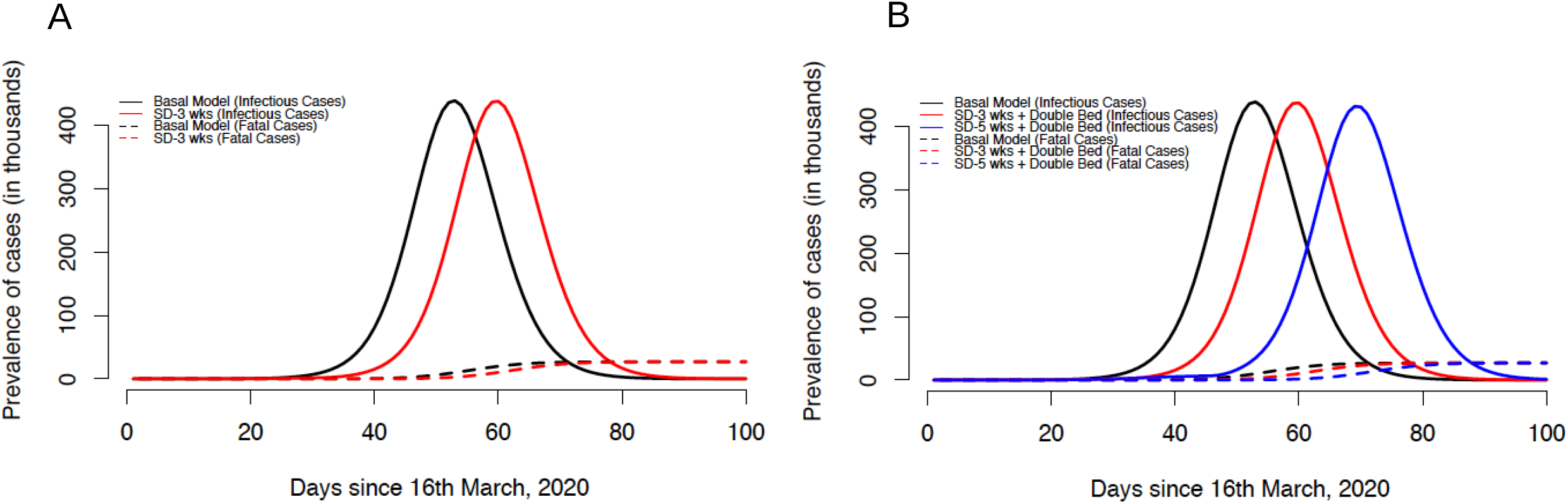
Prevalence of cases in the Infectious and Fatal compartment due to Social Distancing (SD) (A) and increasing hospital bed capacity (B).

### 3.2 Impact of Increased Case detection and Isolation Rate in addition to Social Distancing and Increasing Hospital Beds on the Prevalence of Infection and Fatality

Having intervened in SD and hospital bed capacity, we next tested whether increasing the rate of case detection followed by immediate quarantine had any effect on reducing the peak prevalence. The peak prevalence is drastically reduced on increasing the detection rate by 3 times and further reduced if increased by 5 times (Figure 4A). The reduction in peak prevalence of the Fatal (F) compartment was also similar (Figure 4B). Continuous SD for 5 weeks with a detection rate of 1/5 (1 out of 5 cases daily gets detected and quarantined) has an enormous effect on reducing peak prevalence and also provides enough time window for healthcare capacity building for the upcoming burden of hospitalization. However, such prolonged lock-down would have a significant negative impact on the economy as well as mental well-being. Considering this, we finally tested whether a continuous SD for 5 weeks or SD for 6 weeks with a gap of 1 week is more effective in reducing the burden of cases. Here we saw that the later approach of SD for 3 weeks followed by a release of 1 week and reintroducing SD for 3 weeks again reduced the peak prevalence of Infectious cases by less than a half compared to the baseline model (Figure 4A). This intervention not only reduced the peak burden of both I and F compartment but also delayed it.

**Figure 4:**
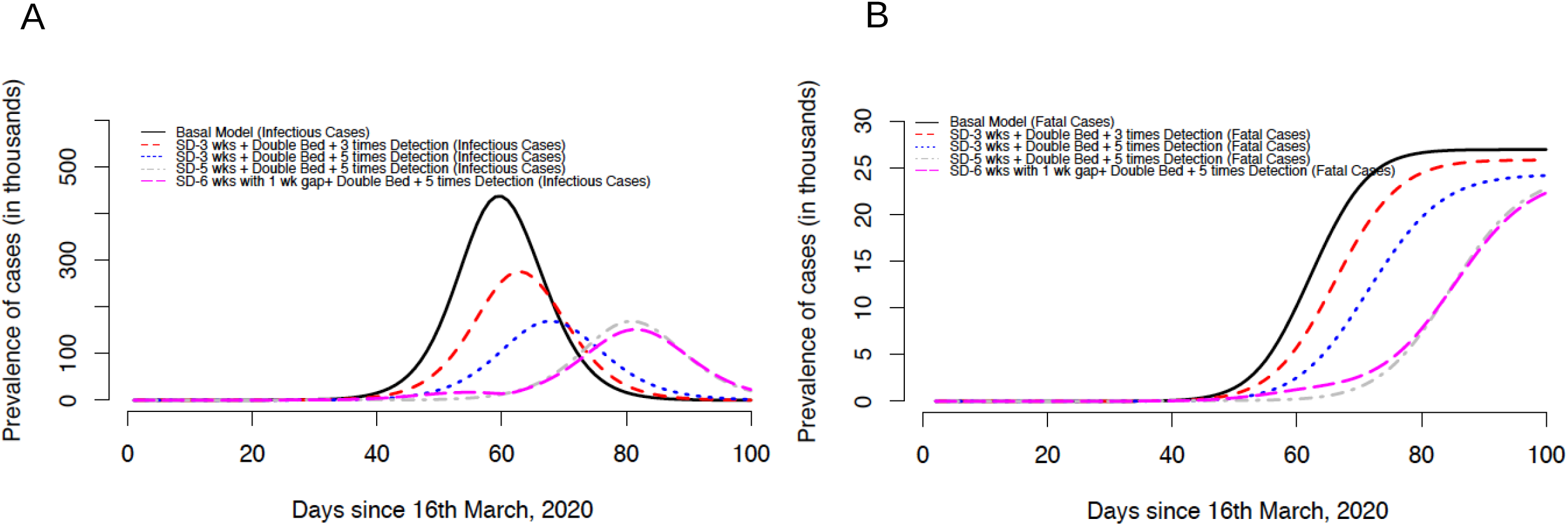
Comparative prevalence of cases in the Infectious (A) and Fatal (B) compartment due to different interventions.

## 4. DISCUSSION

Implementation of Social Distancing or lock-down for 3 weeks has been deployed in India from 25^th^ March, 2020 as an effective measure towards curtailing exposure and the prevalence of infection of COVID-19. These measures are also accompanied by increasing bed capacity to decrease severe as well as the fatal cases. However, for an effective mitigation of an epidemic, detection of unidentified cases with mild and no-symptoms who can expose a large population is of utmost importance. Indeed, a recent study has shown that 86% of all infections were undocumented and thus could be implicated to the rapid geographic spread of COVID-19 [17]. Using a Stochastic Computational Simulation Model, we find that a reduction in Infectious (I) and Fatal (F) COVID-19 cases is possible through increased case detection in addition to implementing SD and increasing the number of hospital beds.

Responding to any epidemic ideally involves the following steps of anticipation, early detection, control & mitigation, containment, elimination or eradication [18]. With the present pandemic of SARS-CoV-2 the global scientific community is still grappling for evidence based, cost effective & sustainable interventions. The best strategy to address the pandemic would only be understood in retrospect, once the storm has blown over. Presently every country is battling, using fundamental process of problem solving ‘trial & error’ method in the middle of the pandemic. Intelligence sharing with quality data management across the countries and globe would help the cause.

Different countries are apparently in different stages of the pandemic; distribution, transmission and outcomes are not uniform for all the countries. Daily confirmed cases vary from 3/million in India to 2816/million population in Spain. Death rates, age and sex distribution, severity pattern are also not same everywhere [19]. The only pattern remaining constant since its inception is the upward curve of the cases in almost all the countries. Given the diabolical nature of the pandemic with prevailing uncertainty over the natural history of the disease and absence of a specific cure or vaccine, panic and anxiety over the disease is ever increasing. In this context do we propose that key to success lies in the detection of cases at the earliest. Even though we do not have a specific pharmacotherapy as yet, non pharmaceutical interventions in addition to that would help to mitigate the situation. Case detection and tracking of the close contacts followed by immediate quarantine measures will eventually help in breaking the chain of transmission.

The extent and intensity of social distancing is in the eye of the storm. The disease propagation follows a somewhat fractal pattern, affecting a cluster of close contacts of a case in a locality and later spreads to other regions. Till the clusters coalesce, disease is localised in smaller localities and that is the best opportunity for our ongoing intervention to work. In South Korea almost 2/3^rd^ of the cases occurred in one particular region of Daegu. While overall incidence is around 19/100000 in the country, Daegu records incidence of 278/100000. Data also shows that only 17% cases were documented as sporadic cases and rest were considered to have occurred in clusters; 50% of that too in one cluster [20]. Hence the spread of the disease is not uniform or homogeneous. Current evidence shows a fall in number of new cases in the recent past in that country. Interestingly, in India also 4 states contribute to about half of all the cases.

Detection of disease is critical. Even for a very highly sensitive and specific test, with low generalised prevalence rate in countries like India, yield of indiscriminate and universal testing would not provide satisfactory result; positive predictive value will be quite low. Naturally the policy makers are justified in setting up criteria for testing based on certain clinical features. However negative test result would practically rule the asymptomatic subject out as infected with SARS-CoV-2 infection.

As the model indicates, a multi-pronged approach of testing, tracking and treating should work. Role of non pharmaceutical measures can never be undermined, especially in this situation. But if one can have a targeted intervention plan centring on a diagnosed case, it will probably be cost effective and sustainable. The social distancing would not only help in restricting the spread, it will provide the much needed leeway, within which we can regroup; and even during the middle of an epidemic we can have the opportunity to strengthen the health system for the upcoming onslaught, if any.

One important limitation of the present study is that the analysis on one geographically confined district may not be implicated to all Indian states and districts with diverse location, population density, availability of healthcare facility, population migration pattern and climatic variations. However, as a proof of concept study we believe our work provides a framework of intervention modalities towards developing policies in mitigating the present epidemic. Moreover, Epidemic Response Planning is needed at district level as inter-district movement become restricted during this period. Our study further suggests that co-ordination of Public and Private Healthcare Sectors would crucial by increasing the hospital beds as well as by supporting medical emergencies.

Towards devising an effective strategy to contain the epidemic spread of COVID-19 in Indian context, our study emphasises the critical importance of increasing detection rates in combination with already existing SD measures and increase in hospital beds. Thus, the best strategy to this end is a combination of three levels of interventions at their optimum values.

## Data Availability

Not Applicable

## Author Contributions

JS, SB, PG and PC contributed to the study concept and design. JS, SP and PG did the data analysis. JS, PG and PC wrote the manuscript. JS, PG and PC are the guarantors of the work. All the authors approved the final version of the article.

## Acknowledgements

JS received a research fellowship from ICMR (No.3/1/3/JRF-2017/HRD-LS/56429/54).

## Disclosure Summary

The authors declare no conflict of interest.

## Notes

### Competing Interest Statement

The authors have declared no competing interest.

